# Maternal smoking during pregnancy and type 1 diabetes in the offspring: A nationwide register-based study with family-based designs

**DOI:** 10.1101/2022.01.07.22268904

**Authors:** Yuxia Wei, Tomas Andersson, Jessica Edstorp, Josefin E. Löfvenborg, Mats Talbäck, Maria Feychting, Sofia Carlsson

## Abstract

**Objectives:** Maternal smoking during pregnancy is associated with a reduced risk of type 1 diabetes (T1D) in the offspring. We investigated whether this association is consistent with a causal interpretation by accounting for familial (shared genetic and environmental) factors using family-based, quasi-experimental designs.

**Design:** A nationwide, prospective cohort study and a nested case-control study (quasi-experiment) comparing children with T1D to their age-matched siblings (or cousins).

**Setting:** Swedish national registers.

**Participants:** We included 2,995,321 children born in Sweden between 1983 and 2014.

**Exposure:** Information on maternal smoking during pregnancy was retrieved from the Swedish Medical Birth Register.

**Main outcome measures:** Children were followed for a diagnosis of T1D until 2020 through the National Patient, Diabetes and Prescribed Drug Registers.

**Results:** A total of 18,617 children developed T1D, with a median age at diagnosis of 9.4 years. The sibling and cousin comparison design included 14,284 and 7,988 of these children, respectively. Maternal smoking during pregnancy was associated with a 22% lower risk of offspring T1D in the full cohort (hazard ratio: 0.78, 95% confidence interval [CI]: 0.75 to 0.82) in the multivariable-adjusted model. The corresponding odds ratio was 0.78 (95% CI: 0.69 to 0.88) in the sibling and 0.72 (95% CI: 0.66 to 0.79) in the cousin comparison analysis.

**Conclusions:** This nationwide, family-based study provides support for a protective effect of maternal smoking on offspring T1D. Mechanistic studies are needed to elucidate the underlying pathways behind this link.

## Introduction

Type 1 diabetes (T1D) is one of the most common chronic diseases in childhood and incidence has been increasing globally over the past three decades at an average annual rate of 3-4% ^1^. Autoantibodies associated with development of T1D may appear already before the age of 6 months but most commonly during the second year of life^2^. This phenomenon indicates that early life factors may contribute to the development of T1D, although few risk factors have been established^2^.

Maternal smoking during pregnancy has been linked to a reduced risk of T1D in the offspring^3^. This association could be due to residual confounding since mothers who smoke during pregnancy may differ from other women in many ways, including child-rearing practices^4 5^ and genetic factors, that may affect offspring risk of T1D but were not adjusted for in most previous studies^3 6-17^. Since it is impossible to perform randomized clinical trials to assess smoking effects on fetal development, other designs are needed to minimize potential confounding.

Family-based designs, such as sibling and cousin comparisons, are quasi-experiments that allows us to control for factors shared by relatives ^18^. E.g., by comparing diabetes risk in siblings who are discordant for fetal exposure to smoking, we can reduce the confounding from genetic (full siblings share 50% of their segregating genes), maternal (intra-uterine and child-rearing), as well as childhood environmental factors. Cousins share fewer familial (genetic and environmental) factors than siblings, by combing cousin and sibling designs we can therefore explore the degree to which genetic and environmental factors account for an observed association^18^.

We aimed to assess the association between maternal smoking during pregnancy and offspring T1D while accounting for familial confounding. To this aim, we used nationwide data from Swedish national registers, and family-based designs. For comparison purposes, we also investigated the association of maternal smoking during pregnancy with offspring type 2 diabetes (T2D).

## Methods

### Registry linkage

Data for this study were retrieved from national registers in Sweden, including the Medical Birth Register (MBR)^19^, the Multi-Generation Register (MGR)^20^, the Longitudinal integrated database for health insurance and labour market studies (LISA)^21^, the National Patient Register (NPR)^22^, the National Diabetes Register (NDR)^23^, the National Prescribed Drug Register (NPDR)^24 25^, and the Total Population Register (TPR)^26^. These registers were linked by the unique personal identity number assigned to every Swedish citizen (**Figure 1**). The study was approved by the Swedish ethical review board (2021-02881).

**Figure 1.**
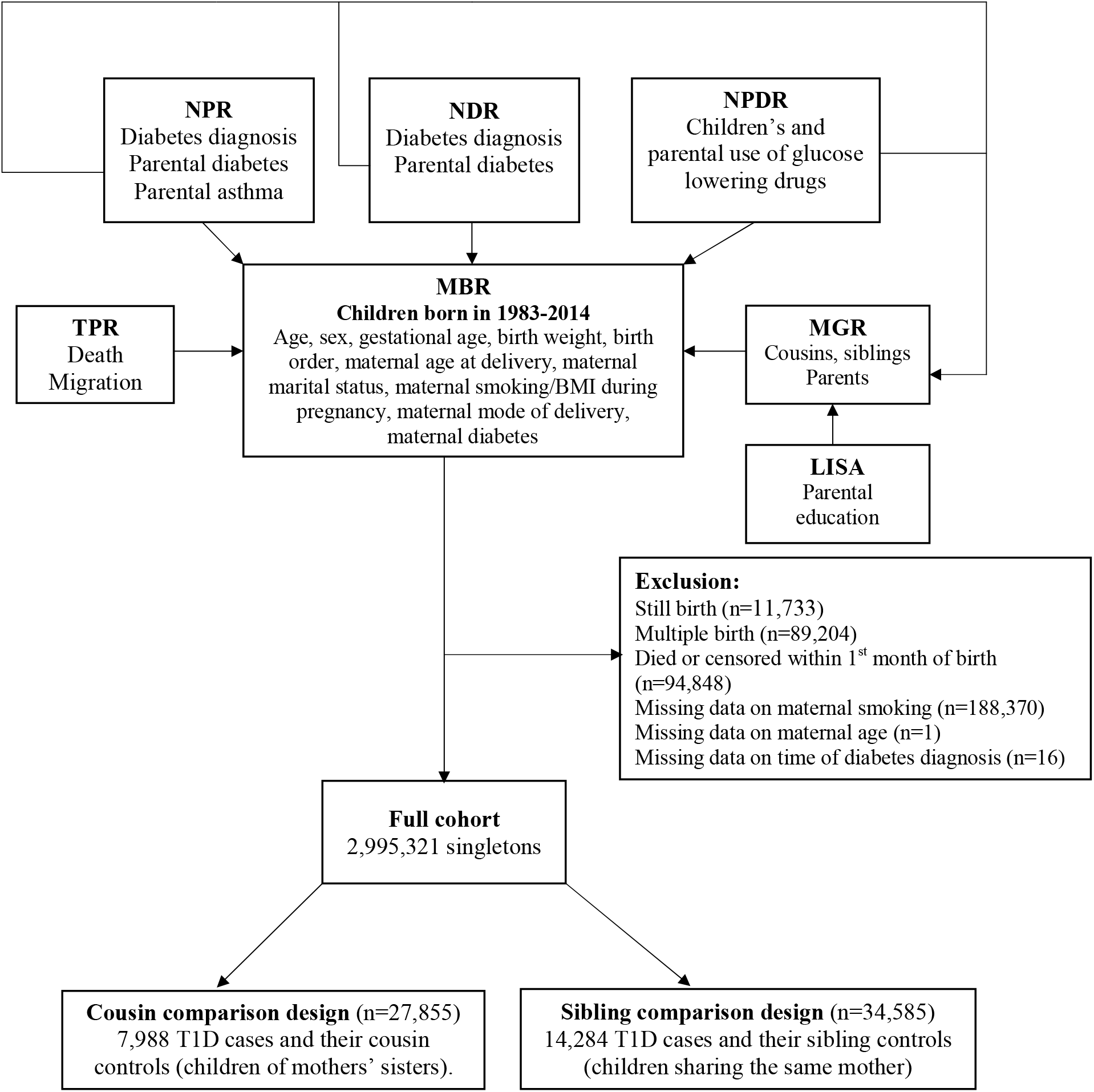
Flow chart of the family-based designs. MBR: the Medical Birth Register; MGR: the Multi-Generation register; LISA: the Longitudinal integrated database for health insurance and labour market studies; TPR: the Total Population Register; NPR: the National Patient Register; NDR: the National Diabetes Register; and NPDR: the National Prescribed Drug Register; T1D: type 1 diabetes.

### Study population

We identified all children born between January 1^st^, 1983, and December 31^st^, 2014 and their mothers through the MBR. We excluded still births (n=11,733), multiple births (n=89,204), children who died or were censored within the first month of birth (n=94,848), individuals failing to be linked to the MGR for relatives’ information (n=39,304), individuals with missing data on maternal smoking during early pregnancy (n=188,370) or maternal age at delivery (n=1) or time of diabetes diagnosis (n=16). This left an analytical sample consisting of 2,995,321 children (**Figure 1**).

### Smoking

Information on maternal smoking during pregnancy was retrieved from MBR, which contains self-reported smoking information since 1983^19^. Trained midwives collected information on smoking from expectant women at their first antenatal visit (typically at 8-12 weeks of pregnancy) using standardized questionnaires. Available response options included non-smoking, 1-9 cigarettes per day, and ≥10 cigarettes per day^27^. A previous study showed that validity of self-reported smoking information during early pregnancy is high in Sweden^28^. MBR also includes information on smoking at 30-32 weeks of pregnancy since 1990, but the missing rate was 75.2% in 1990s, 24% in 2000s, and 12.7% in 2010s. Unless stated otherwise, maternal smoking during pregnancy refers to maternal smoking at the first antenatal visit in the following sections.

### Diabetes

The children were followed for a diagnosis of T1D at age ≤18 years by linkage to NPR, NDR, and NPDR. NPR was established in 1964 and covers all inpatient care in Sweden since 1987 and outpatient specialist care provided by public and private caregivers since 2001^22^. NPR coded diseases according to International Classification of Diseases (ICD). NDR was created in 1996 and is the largest diabetes register in the world (www.ndr.nu)^23^. NPDR was established in July 2005 and includes all prescribed drugs dispensed at Swedish pharmacies, with prescribed drugs coded by the Anatomical Therapeutic Code (ATC)^24^.

T1D was defined as receiving a diagnosis of T1D at age ≤ 18 years (a) in NPR (ICD-8 code 250, ICD-9 code 250, ICD-10 code E10) or (b) in NDR, or c) the exclusive use of insulin at age≤18 years recorded in NPDR. Patients were also defined as T1D if they were diagnosed with diabetes of unknown type or of different types before age of 18 years in NPR or NDR and did not have a record of oral glucose lowering drugs in NPDR.

We also identified cases of T2D in these registers as a secondary outcome. T2D was defined as receiving a diagnosis of T2D (a) in NPR (ICD-10 code E11) or (b) in NDR, with or without the use of glucose lowering drugs. Patients were also defined as T2D if they were diagnosed with diabetes of unknown type in NPR or NDR but were exclusively prescribed with non-insulin glucose lowering drugs.

Date of diagnosis was defined according to the first recording in NDR, NPR or NPDR, whichever came first. In addition, we checked vital status and migration status of participants by linking to TPR^26^.

### Covariates

MBR contains information on year and month of birth, sex, gestational age, birth weight, birth order, maternal age at delivery, maternal marital status, maternal body mass index (BMI) at first antenatal visit, and mode of delivery. Data on parental highest degree of education attained before childbirth were obtained from LISA^21^. Parental diabetes at childbirth was identified from NDR, NPR and NPDR. Parental asthma at childbirth was identified from the NPR.

### Family-based, nested case-control study

Within the study population, we linked siblings and cousins to each other through their parents and grandparents in MGR, which contains information on parents of children born from 1932 onwards^20^. In the sibling comparison design, children with T1D were matched to their siblings (same mother), who were alive and free of diabetes at the age when the case was diagnosed^29^. In the cousin comparison design, we only included the offspring of sisters and matched all children with T1D to their cousins who were alive and free of diabetes at the age when the case was diagnosed. Sibling (or cousin) groups that were discordant on both maternal smoking and T1D diagnosis contributed to the estimates for maternal smoking during pregnancy^29 30^. However, sibling (or cousin) groups concordant on maternal smoking were informative for the estimates for covariates and were therefore included as participants^29^. The same approach of matching was used for the cousin and sibling comparison analyses of T2D.

### Statistical analysis

#### Cohort analysis

For the analysis of T1D, follow-up time was calculated from the date of birth to the occurrence of diabetes, death, migration, December 31^st^ of the year when children turned18 years old, or December 31^st^, 2020, whichever came first. Analyses were similar for T2D but without censoring for age. Cox proportional hazards regression models estimated the hazard ratios (HRs) and 95% confidence intervals (CIs) for maternal smoking during pregnancy. The models were fitted with age as the time scale and a gamma-frailty component^31^ to account for familial clustering of participants.

#### Sibling and cousin comparison analyses

In the sibling (or cousin) comparison analysis, conditional logistic models estimated the odds ratios (ORs) and 95% CIs of T1D and T2D for maternal smoking during pregnancy, conditioning on sibling (or cousin) groups.

Models in the cohort, sibling and cousin analyses were all adjusted for sex, year of birth (model 1), maternal and paternal education, maternal age at delivery, maternal BMI at first antenatal visit, maternal diabetes status at childbirth, paternal diabetes at childbirth, birth order (model 2), gestational age, and birth weight for gestational age (model 3). Participants with missing values on categorical covariates were treated as a separate group in the analyses and those with missing values on continuous covariates were assigned the median value. A binary variable was included in the analyses to indicate if values were imputed.

#### Timing of maternal smoking during pregnancy

We also explored the incidence of offspring T1D in relation to maternal smoking only at first antenatal visit, only at 30-32 weeks of pregnancy, and sustained smoking during pregnancy (both at first antenatal visit and at 30-32 weeks of pregnancy) in the subset of children (n=1,363,501) born after 2000 when such information was recorded with a relatively low missing rate.

#### Sensitivity analyses

We did subgroup analyses according to sex and birth year. To assess the generalizability of the sibling results, the cohort analyses were performed separately in individuals with and without siblings.

We also ran the analyses with additional adjustment for maternal marital status, mode of delivery and parental history of asthma (to account for parental asthma’s potential influence on maternal smoking and the shared genetic susceptibility between asthma and T1D^32 33^), and performed complete-case analysis where individuals with missing data on covariates were excluded. We limited the sibling comparison analyses to full siblings, and the cousin comparison analyses to full cousins (offspring of sisters who were full siblings). We also restricted the sibling (cousin) analyses to siblings (cousins) born within 5 years, who may share childhood environmental factors to a larger extent than those with a larger age difference. The sibling comparison design assumes no carryover effect, that is, the first sibling’s maternal smoking during pregnancy should not affect the subsequent sibling’s risk of T1D^34-36^. The carryover effect is less of a concern in the cousin comparison design^36^. To further minimize the potential bias from a carryover effect, we limited the cousin comparison analysis to first-born cousins. To assess the potential for unmeasured confounding, we calculated an E value^37^ (**eMethods**) based on the OR of T1D associated with maternal smoking from the sibling analysis (model 3).

The gamma-frailty models were performed in R 4.1.0 and other statistical analyses were performed in Stata 16.1.

### Patient and Public Involvement

Neither participants nor the public were involved in setting the research question or the outcome measures, nor were they involved in the study design and result interpretation. The results will be disseminated to members of the public through appropriate media channels.

## Results

### Characteristics

A total of 18,617 T1D cases diagnosed at age ≤18 years occurred during a median follow-up of 18.2 years in the cohort of 2,995,321 individuals. The median age at T1D diagnosis was 9.4 years. The cousin comparison included 7,988 T1D cases who had at least one (range: 1-26; median: 3) eligible cousin (n=27,855) and the sibling analyses included 14,284 T1D with at least one (range: 1-10; median: 1) eligible sibling (n= 34,585) (**Figure 1**). Compared to children without T1D, cases were more likely to be boys and have parents with diabetes (**Table 1**). Children whose mothers smoked during pregnancy were more likely to be small for gestational age, have younger mothers, and have parents with lower levels of education (**eTable 1**).

**Table 1.** Characteristics of children by T1D status in different study designs^a^.

|  | Cohort analysis |  | Cousin analysis |  | Sibling analysis |  |
| --- | --- | --- | --- | --- | --- | --- |
|  | No T1D | T1D | No T1D | T1D | No T1D | T1D |
| Boys | 1,529,498 (51.4) | 10,166 (54.6) | 10,729 (51.0) | 4,399 (55.1) | 10,908 (51.2) | 7,816 (54.7) |
| Year of birth |  |  |  |  |  |  |
| 1980-1984 | 164,968 (5.5) | 888 (4.8) | 1,141 (5.4) | 302 (3.8) | 799 (3.8) | 451 (3.2) |
| 1985-1989 | 469,384 (15.8) | 2,811 (15.1) | 3,800 (18.1) | 12,203 (15.1) | 3,537 (16.6) | 2,053 (14.4) |
| 1990-1994 | 534,392 (18.0) | 3,892 (20.9) | 4,918 (23.4) | 1,849 (23.1) | 5,038 (23.7) | 3,236 (22.7) |
| 1995-1999 | 407,468 (13.7) | 3,404 (18.3) | 3,769 (17.9) | 1,476 (18.5) | 4,192 (19.7) | 2,790 (19.5) |
| 2000-2004 | 421,330 (14.2) | 3,533 (19.0) | 3,436 (16.3) | 1,531 (19.2) | 3,782 (17.8) | 2,842 (19.9) |
| 2005-2009 | 468,632 (15.7) | 2,696 (14.5) | 2,624 (12.5) | 1,090 (13.6) | 2,656 (12.5) | 2,032 (14.2) |
| 2010-2014 | 510,530 (17.2) | 1,393 (7.5) | 1,348 (6.4) | 537 (6.7) | 1,298 (6.1) | 80 (6.2) |
| Gestational age in days,<br>mean (SD) | 278.8 (12.2) | 277.9 (12.2) | 278.4 (12.4) | 278.0 (12.0) | 278.3 (12.0) | 278.0 (12.0) |
| Birth weight for<br>gestational age |  |  |  |  |  |  |
| Small for gestational age | 70,763 (2.4) | 382 (2.1) | 466 (2.2) | 167 (2.1) | 388 (1.8) | 267 (1.9) |
| Normal | 2,793,435 (93.8) | 17,350 (93.2) | 19,663 (93.5) | 7,466 (93.5) | 19,783 (92.9) | 13,316 (93.2) |
| Large for gestational age | 103,661 (3.5) | 834 (4.5) | 850 (4.0) | 337 (4.2) | 1,066 (5.0) | 666 (4.7) |
| Unknown | 8,845 (0.3) | 51 (0.3) | 57 (0.3) | 18 (0.2) | 65 (0.3) | 35 (0.2) |
| Birth order |  |  |  |  |  |  |
| 1 | 1,265,549 (42.5) | 7,851 (42.2) | 8,667 (41.2) | 3,273 (41.0) | 7,523 (35.3) | 5,203 (36.4) |
| 2 | 1,082,395 (36.4) | 6,833 (36.7) | 7,545 (35.9) | 2,966 (37.1) | 7,617 (35.8) | 5,796 (40.6) |
| 3+ | 628,760 (21.1) | 3,933 (21.1) | 4,824 (22.9) | 1,749 (21.9) | 6,162 (28.9) | 3,285 (23.0) |
| Maternal age at delivery<br>in years, mean (SD) | 29.7 (5.2) | 29.7 (5.1) | 28.8 (5.1) | 29.4 (5.1) | 29.0 (5.1) | 29.5 (5.0) |
| Maternal marital status |  |  |  |  |  |  |
| Living with child's father | 2,751,651 (92.4) | 17,282 (92.8) | 19,354 (92.0) | 7,420 (92.9) | 19,788 (92.9) | 13,362 (93.5) |
| Single | 56,402 (1.9) | 330 (1.8) | 452 (2.1) | 151 (1.9) | 347 (1.6) | 227 (1.6) |
| Other | 89,494 (3.0) | 482 (2.6) | 641 (3.0) | 196 (2.5) | 553 (2.6) | 293 (2.1) |
| Unknown | 79,157 (2.7) | 523 (2.8) | 589 (2.8) | 221 (2.8) | 614 (2.9) | 402 (2.8) |
| Maternal educational level |  |  |  |  |  |  |
| Pre-secondary | 386,663 (13.0) | 2,343 (12.6) | 2,799 (13.3) | 980 (12.3) | 2,705 (12.7) | 1,600 (11.2) |
| Upper-secondary | 567,201 (19.1) | 4,343 (23.3) | 5,282 (25.1) | 2,105 (26.4) | 5,108 (24.0) | 3,459 (24.2) |
| High school | 567,452 (19.1) | 3,573 (19.2) | 3,684 (17.5) | 1,495 (18.7) | 3,764 (17.7) | 2,736 (19.2) |
| Post-secondary or higher | 914,211 (30.7) | 5,387 (28.9) | 5,018 (23.9) | 2,127 (26.6) | 5,547 (26.0) | 4,198 (29.4) |
| Unknown | 541,177 (18.2) | 2,971 (16.0) | 4,253 (20.2) | 1,281 (16.0) | 4,178 (19.6) | 2,291 (16.0) |
| Paternal educational level |  |  |  |  |  |  |
| Pre-secondary | 492,235 (16.5) | 2,975 (16.0) | 3,597 (17.1) | 1,258 (15.7) | 3,213 (15.1) | 2,098 (14.7) |
| Upper-secondary | 716,097 (24.1) | 5,451 (29.3) | 62,200 (29.5) | 2,491 (31.2) | 6,296 (29.6) | 4,334 (30.3) |
| High school | 547,304 (18.4) | 3,226 (17.3) | 3,287 (15.6) | 1,344 (16.8) | 3,312 (15.5) | 2,372 (16.6) |
| Post-secondary or higher | 785,763 (26.4) | 4,607 (24.7) | 4,347 (20.7) | 1,828 (22.9) | 5,002 (23.5) | 3,645 (25.5) |
| Unknown | 435,305 (14.6) | 2,358 (12.7) | 3,605 (17.1) | 1,067 (13.4) | 3,479 (16.3) | 1,835 (12.8) |
| Maternal BMI in kg/m<sup>2</sup><br>during pregnancy<sup>b</sup>, mean (SD) | 23.9 (4.3) | 24.1 (4.3) | 23.9 (4.3) | 24.2 (4.3) | 24.1 (4.5) | 24.1 (4.3) |
| Maternal caesarean section<br>at delivery | 399,662 (13.4) | 2,724 (14.6) | 2,595 (12.3) | 1,107 (13.9) | 2,576 (12.1) | 1,890 (13.2) |
| Maternal diabetes at childbirth | 24,923 (0.8) | 593 (3.2) | 340 (1.6) | 256 (3.2) | 560 (2.6) | 392 (2.7) |
| Paternal diabetes at childbirth | 21,731 (0.7) | 865 (4.6) | 123 (0.6) | 371 (4.6) | 760 (3.6) | 623 (4.4) |
| Maternal asthma at childbirth | 158,268 (5.3) | 1,085 (5.8) | 1,093 (5.2) | 475 (5.9) | 1,141 (5.4) | 815 (5.7) |
| Paternal asthma at childbirth | 35,258 (1.2) | 237 (1.3) | 202 (1.0) | 114 (1.4) | 259 (1.2) | 171 (1.2) |

T1D: type 1 diabetes, SD: standard deviation; BMI: body mass index.
<sup>a</sup> Values are presented as numbers (proportions [%]) unless stated otherwise.<sup>b</sup> At first antenatal visit, mostly during 8-12 weeks' of pregnancy.

### Maternal smoking during pregnancy and offspring T1D

In the cohort analysis, the HR (95% CI) of offspring T1D was 0.78 (0.75 to 0.82) for maternal smoking versus nonsmoking during pregnancy (**Figure 2**). The inverse association was similar in the cousin (OR: 0.72, 95% CI: 0.66 to 0.79) and the sibling comparison analysis (OR: 0.78, 95% CI: 0.69 to 0.88) (**Figure 2**). The associations did not change across three models.

**Figure 2.**
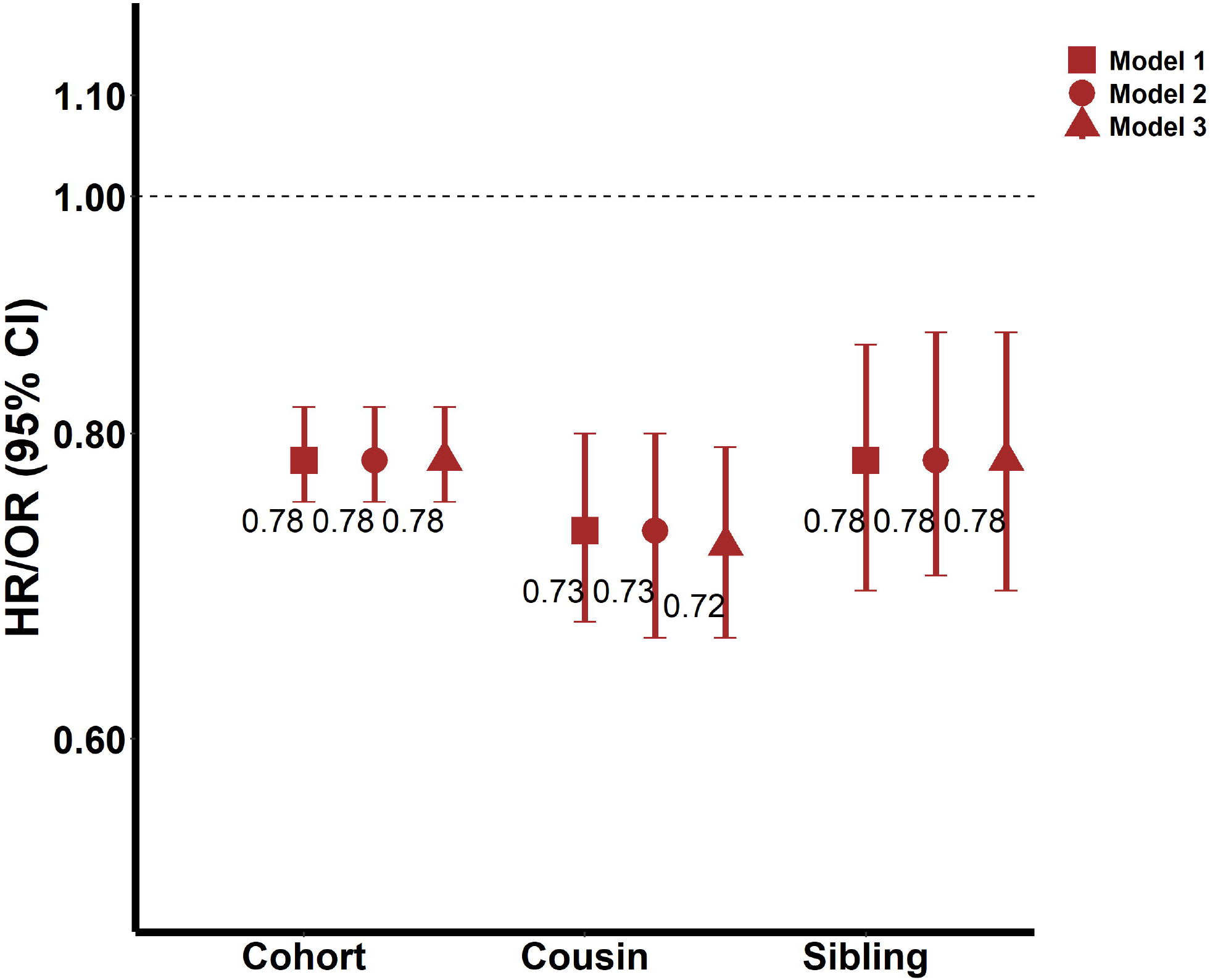
HRs/ORs (95% CIs) of T1D for maternal smoking versus nonsmoking during pregnancy in different study designs. T1D: type 1 diabetes; HR: hazard ratio; OR: odds ratio; CI: confidence interval Model 1 was adjusted for sex and year of birth. The model in the cohort analysis was fitted with a frailty component to take into account the nonindependence among children born by the same mother. The model in the cousin (or sibling) analysis was conditioning on cousin (or sibling) groups. Model 2 was additionally adjusted for maternal education, paternal education, maternal age at delivery, maternal body mass index during pregnancy, maternal diabetes, paternal diabetes, and birth order on the basis of model 1. Model 3 was additionally adjusted for gestational age and birth weight for gestational age on the basis of model 2.

Both smoking 1-9 cigarettes/day and ≥10 cigarettes/day were associated with a reduced risk of T1D in the offspring (**Table 2**), with an OR of 0.80 (95% CI: 0.71 to 0.90) and 0.71 (95% CI: 0.60 to 0.84), respectively in the sibling analyses (**Table 2**).

**Table 2.** Associations between maternal smoking during pregnancy and offspring T1D in different study designs.

| **Smoking status** | **Nonsmoking** | **1-9 cigarettes/day** | **≥10 cigarettes/day** | **P for test of trend** |
| --- | --- | --- | --- | --- |
| **Cohort analysis** |  |  |  |  |
| No. of cases | 16,084 | 1,684 | 849 |  |
| Person years | 38,819,013 | 5,235,610 | 2,863,183 |  |
| Incidence (no./10,000 person years) | 4.1 | 3.2 | 3.0 |  |
| Model 1 | 1.00 | 0.80 (0.76 to 0.85) | 0.75 (0.70 to 0.80) | 0.100 |
| Model 2 | 1.00 | 0.80 (0.76 to 0.84) | 0.74 (0.69 to 0.80) | 0.076 |
| Model 3 | 1.00 | 0.80 (0.76 to 0.85) | 0.74 (0.69 to 0.80) | 0.066 |
| **Cousin comparison analysis** |  |  |  |  |
| No. of cases | 6,765 | 801 | 422 |  |
| Model 1 | 1.00 | 0.75 (0.68 to 0.83) | 0.69 (0.60 to 0.79) | 0.245 |
| Model 2 | 1.00 | 0.75 (0.68 to 0.83) | 0.67 (0.58 to 0.77) | 0.114 |
| Model 3 | 1.00 | 0.75 (0.68 to 0.83) | 0.66 (0.58 to 0.76) | 0.111 |
| **Sibling comparison analysis** |  |  |  |  |
| No. of cases | 12,483 | 1,193 | 608 |  |
| Model 1 | 1.00 | 0.79 (0.71 to 0.89) | 0.70 (0.60 to 0.83) | 0.116 |
| Model 2 | 1.00 | 0.80 (0.71 to 0.90) | 0.71 (0.60 to 0.84) | 0.119 |
| Model 3 | 1.00 | 0.80 (0.71 to 0.90) | 0.71 (0.60 to 0.84) | 0.115 |

T1D: type 1 diabetes
Model 1 was adjusted for sex and year of birth. The Cox model in the cohort analysis was fitted with a gamma-frailty component to account for the clustering of siblings from the same mother. The conditional logistic model in the sibling (or cousin) comparison analysis conditioned on sibling (or cousin) groups.
Model 2 was additionally adjusted for maternal education, paternal education, maternal age at delivery, maternal body mass index during pregnancy, maternal history of diabetes, paternal history of diabetes, and birth order based on model 1.
Model 3 was additionally adjusted for gestational age and birth weight for gestational age based on model 2.

In the subset born 2000 onwards with information on timing of smoking, cohort analyses revealed a reduced risk primarily in children of mothers who smoked persistently throughout the pregnancy (HR: 0.83, 95% CI: 0.74 to 0.93). In sibling analyses, numbers were small and the risk reduction appeared similar across the exposure windows, i.e. OR (95% CI) was 0.81 (0.55 to 1.19), 0.66 (0.36 to 1.21) and 0.74 (0.48 to 1.13), respectively for smoking only at first antenatal visit, only at 30-32 weeks of pregnancy, and sustained throughout the pregnancy (**eTable 2**).

### Maternal smoking during pregnancy and offspring T2D

A total of 3,477 T2D cases (median age at diagnosis 26.5 years) occurred during a median follow-up of 21.5 years. There was a positive association between maternal smoking and T2D in the cohort analysis (HR: 1.90, 95% CI: 1.77 to 2.04) which was attenuated in the cousin comparison analysis (OR: 1.43, 95% CI: 1.18 to 1.73) and even more so in the sibling analysis (OR: 1.23, 95% CI: 0.93 to 1.62) (**eFigure 1**).

### Sensitivity analyses

The association between maternal smoking during pregnancy and offspring T1D was similar when cohort analyses were performed separately in participants with and without siblings (**eTable 3**). Sex and year of birth did not seem to modify the associations in neither cohort (**eTable 3**) nor cousin or sibling comparison analyses (**eTable 4**). Results were similar after additional adjustment for maternal marital status, mode of delivery or parental asthma at childbirth, and by exclusion of individuals with missing data on any covariate (**eTable 5-7**). Similarly, associations remained in cousin analyses restricted to full cousins, cousins born within 5 years, and first-born cousins (**eTable 6**), as well as in sibling analyses restricted to full siblings and siblings within 5 years of age difference (**eTable 7**). The E value was 1.88 calculated based the estimated OR in model 3 of the sibling comparison analysis, which controlled for the most unmeasured familial confounders.

## Discussion

### Main findings

This nationwide prospective study shows that children exposed to maternal smoking during pregnancy have a 22% lower risk of developing T1D during childhood compared to their unexposed siblings. This confirms results of previous observational studies^3^ and suggests that maternal smoking during pregnancy has the potential to prevent offspring T1D.

### Main findings in relation to previous studies

We integrated cohort, sibling and cousin designs, the latter two of which allowed us to control for potential unmeasured confounders shared within families. The observed association between maternal smoking during pregnancy and offspring T1D was consistent across the three designs, and the strength of the association was in line with the RR of 0.72, observed in a recent meta-analysis based on five cohort studies^3^. This indicates that the association is unlikely to be explained by confounding and furthermore, that the influence of familial (genetic and early environmental) factors is minor. According to our estimated E value, an uncontrolled confounder needs to have a risk ratio of 1.88 with both the exposure and outcome to fully explain away the inverse association between maternal smoking during pregnancy and offspring T1D. It should be noted that we had the opportunity to adjust for both unmeasured familial factors and a range of measured perinatal factors including mode of delivery, maternal BMI, parental diabetes, and gestational age, and we are not aware of any other factor with such a strong association to both maternal smoking and T1D.

With regard to the timing of maternal smoking during pregnancy, results of previous studies are inconsistent^3 7 38^. Our cohort analysis found a reduced risk primarily in women who continued smoking throughout the pregnancy. However, the results of sibling analyses, although based on small numbers, were consistent with a reduced risk also in the offspring of women who quit smoking after the first trimester.

There was a positive association between offspring T2D and maternal smoking during pregnancy in the cohort analyses, which was attenuated in the sibling comparison analysis. This suggests that familial confounding contributes to the association observed in the full cohort. These findings are in line with those of a previous meta-analysis of five studies^39^ showing no association between maternal smoking during pregnancy and offspring T2D.

### Potential mechanisms

The inverse association between maternal smoking during pregnancy and offspring T1D could hypothetically be attributed to immunosuppressive effects of nicotine. Experimental studies show that nicotine can activate nicotinic acetylcholine receptors (nAChRs) in T cells^40 41^. The activation of nAChRs may subsequently suppress systemic inflammation and autoimmunity^40 41^. Nicotine induced immunosuppression is associated with preserved insulin content and reduced incidence of diabetes in T1D-prone mice models^42^. Experimental studies indicate that the immunosuppressive and anti-inflammatory effects of prenatal smoking exposure remain also postnatally^43 44^. The hypothesis of immunosuppression is partly supported by the fact that autoantibodies of T1D mostly appear within the first two years after birth, close to the time of fetal exposure^2^. In this context it is noteworthy that parental smoking during childhood appears unrelated to offspring T1D risk^38^, supporting that the fetal period may be a sensitive period for a proposed smoking effect. The lack of association observed between maternal smoking and T2D supports that the mechanism may involve effects related primarily to autoimmunity rather than other diabetogenic processes. Future mechanistic studies are however warranted to elucidate the potential pathways linking maternal smoking to offspring risk of T1D. While these results do not have immediate clinical relevance, they do provide insights into the etiology and pathogenesis of T1D.

### Strength and limitations

Strengths include the use of nationwide data, with almost 3 million children of whom 18,617 developed T1D. Follow-up was made through the combination of national patient, drug and diabetes registers to ensure a high coverage of diabetes cases and virtually no loss to follow-up. The quality of the registers is high^22 23^; a diagnosis of T1D based on the NDR has been shown to be accurate in 97% of cases diagnosed at age ≤30^23^. A particular strength is the integration of a traditional cohort design with family-based designs of sibling and cousin comparison analyses; this allows us to control for unmeasured environmental and genetic confounders shared within families. In addition, we could adjust for a range of perinatal factors based on information recorded at birth. The possibility to compare results for T1D vs. T2D, provides clues to the potential mechanism linking maternal smoking to offspring T1D. Limitations include self-reported information on maternal smoking during pregnancy which may be underreported due to social desirability. Because this was a prospective study, we can assume misclassification to be non-differential which leads to dilution rather than overestimation of associations. Furthermore, a previous Swedish study indicated that validity of information on maternal smoking during early pregnancy is high^45^. We lacked information on breastfeeding, which has been linked to a reduced risk of offspring T1D^46^. Such potential confounding is most likely reduced by comparing siblings. It should also be noted that since mothers who smoke can be expected to breastfeed less, failure to adjust for breastfeeding is unlikely to explain the inverse association between smoking and T1D. Finally, we lacked information on paternal smoking during pregnancy and parental smoking during childhood. Still, a previous large cohort study did not observe an association between either parental smoking during childhood or paternal smoking during maternal pregnancy and offspring T1D^38^.

## Conclusions

This study provides evidence from family-based designs of sibling and cousin comparison analyses that maternal smoking during pregnancy may have a protective effect on offspring T1D, adding evidence to current knowledge on the development of T1D. Despite these findings, smoking during pregnancy should be strongly advised against since it has several severe harmful effects on fetal and childhood health^30 45 47^.

### Summary boxes

**What is already known for this topic?**

- Previous observational studies have reported a reduced risk of type 1 diabetes (T1D) in the offspring in relation to maternal smoking during pregnancy.
- This association could be due to confounding since mothers who smoke during pregnancy may differ from other women in many ways that are difficult to adjust for.

**What this study adds?**

- This nationwide study is the first study of maternal smoking and T1D that uses family-based, quasi-experimental designs to adjust for genetic and environmental factors shared within families.
- The results show that children prenatally exposed to cigarette smoking have a 22% reduced risk of T1D compared to their unexposed siblings.
- This study provides strong support for a reduced risk of childhood T1D conferred by maternal smoking during pregnancy.

## Data Availability

The data that support the findings of this study are available from Statistics Sweden and the Swedish National Board of Health and Welfare but restrictions apply to the availability of these data, which were used under license for the current study, and so are not publicly available.

## Authors’ contributions

SC and YW conceived and designed the study. MF and MT collected the data. YW analyzed the data and drafted the manuscript. TA contributed to methodological issues. All authors contributed to the interpretation of the results and critically revised the manuscript for valuable intellectual content. All authors reviewed and approved the final manuscript.

## Transparency statement

The lead author affirms that the manuscript is an honest, accurate, and transparent account of the study being reported; that no important aspects of the study have been omitted; and that any discrepancies from the study as originally planned have been explained.

## References

1. Norris JM, Johnson RK, Stene LC. Type 1 diabetes-early life origins and changing epidemiology. Lancet Diabetes Endocrinol 2020;8(3):226–38. doi: 10.1016/s2213-8587(19)30412-7 [published Online First: 2020/01/31]

2. Knip M, Luopajärvi K, Härkönen T. Early life origin of type 1 diabetes. Seminars in Immunopathology 2017;39(6):653–67. doi: 10.1007/s00281-017-0665-6

3. Begum M, Pilkington RM, Chittleborough CR, et al. Effect of maternal smoking during pregnancy on childhood type 1 diabetes: a whole-of-population study. Diabetologia 2020;63(6):1162–73. doi: 10.1007/s00125-020-05111-w [published Online First: 2020/02/26]

4. Rogers I, Emmett P. The effect of maternal smoking status, educational level and age on food and nutrient intakes in preschool children: results from the Avon Longitudinal Study of Parents and Children. Eur J Clin Nutr 2003;57(7):854–64. doi: 10.1038/sj.ejcn.1601619 [published Online First: 2003/06/25]

5. Godleski SA, Shisler S, Eiden RD, et al. Maternal Smoking and Psychosocial Functioning: Impact on Subsequent Breastfeeding Practices. Breastfeed Med 2020;15(4):246–53. doi: 10.1089/bfm.2019.0148 [published Online First: 2020/03/10]

6. Norrman E, Petzold M, Clausen TD, et al. Type 1 diabetes in children born after assisted reproductive technology: a register-based national cohort study. Hum Reprod 2020;35(1):221–31. doi: 10.1093/humrep/dez227 [published Online First: 2020/01/25]

7. Metsälä J, Hakola L, Lundqvist A, et al. Perinatal factors and the risk of type 1 diabetes in childhood and adolescence-A register-based case-cohort study in Finland, years 1987 to 2009. Pediatr Diabetes 2020;21(4):586–96. doi: 10.1111/pedi.12994 [published Online First: 2020/02/01]

8. Ayati M, Mosayebi Z, Movahedian AH. Association Between Perinatal Risk Factors and Development of Type 1 Diabetes in Children. Journal of Comprehensive Pediatrics 2020;11(2) doi: 10.5812/compreped.82902

9. Boljat A, Gunjača I, Konstantinović I, et al. Environmental Risk Factors for Type 1 Diabetes Mellitus Development. Exp Clin Endocrinol Diabet 2017;125(8):563–70. doi: 10.1055/s-0043-109000

10. Hussen HI, Persson M, Moradi T. Maternal overweight and obesity are associated with increased risk of type 1 diabetes in offspring of parents without diabetes regardless of ethnicity. Diabetologia 2015;58(7):1464–73. doi: 10.1007/s00125-015-3580-1 [published Online First: 2015/05/06]

11. Haynes A, Cooper MN, Bower C, et al. Maternal smoking during pregnancy and the risk of childhood type 1 diabetes in Western Australia. Diabetologia 2014;57(3):469–72. doi: 10.1007/s00125-013-3122-7 [published Online First: 2013/12/04]

12. Robertson L, Harrild K. Maternal and neonatal risk factors for childhood type 1 diabetes: a matched case-control study. BMC Public Health 2010;10:281. doi: 10.1186/1471-2458-10-281 [published Online First: 2010/05/29]

13. D’Angeli MA, Merzon E, Valbuena LF, et al. Environmental Factors Associated With Childhood-Onset Type 1 Diabetes Mellitus: An Exploration of the Hygiene and Overload Hypotheses. Arch Pediatr Adolesc Med 2010;164(8):732–38. doi: 10.1001/archpediatrics.2010.115

14. Toschke AM, Ehlin A, Koletzko B, et al. Paternal smoking is associated with a decreased prevalence of type 1 diabetes mellitus among offspring in two national British birth cohort studies (NCDS and BCS70). J Perinat Med 2007;35(1):43–7. doi: 10.1515/jpm.2007.006 [published Online First: 2007/02/23]

15. Ievins R, Roberts SE, Goldacre MJ. Perinatal factors associated with subsequent diabetes mellitus in the child: record linkage study. Diabet Med 2007;24(6):664–70. doi: 10.1111/j.1464-5491.2007.02147.x [published Online First: 2007/04/04]

16. Svensson J, Carstensen B, Mortensen HB, et al. Early childhood risk factors associated with type 1 diabetes--is gender important? Eur J Epidemiol 2005;20(5):429–34. doi: 10.1007/s10654-005-0878-1 [published Online First: 2005/08/06]

17. Dahlquist G, Kallen B. MATERNAL-CHILD BLOOD-GROUP INCOMPATIBILITY AND OTHER PERINATAL EVENTS INCREASE THE RISK FOR EARLY-ONSET TYPE-1 (INSULIN-DEPENDENT) DIABETES-MELLITUS. Diabetologia 1992;35(7):671–75. doi: 10.1007/bf00400261

18. D’Onofrio BM, Lahey BB, Turkheimer E, et al. Critical need for family-based, quasi-experimental designs in integrating genetic and social science research. Am J Public Health 2013;103 Suppl 1(Suppl 1):S46-55. doi: 10.2105/ajph.2013.301252 [published Online First: 2013/08/10]

19. Cnattingius S, Ericson A, Gunnarskog J, et al. A quality study of a medical birth registry. Scand J Soc Med 1990;18(2):143–8. doi: 10.1177/140349489001800209 [published Online First: 1990/06/01]

20. Ekbom A. The Swedish Multi-generation Register. Methods Mol Biol 2011;675:215–20. doi: 10.1007/978-1-59745-423-0_10 [published Online First: 2010/10/16]

21. Ludvigsson JF, Svedberg P, Olén O, et al. The longitudinal integrated database for health insurance and labour market studies (LISA) and its use in medical research. Eur J Epidemiol 2019;34(4):423–37. doi: 10.1007/s10654-019-00511-8 [published Online First: 2019/04/01]

22. Ludvigsson JF, Andersson E, Ekbom A, et al. External review and validation of the Swedish national inpatient register. BMC Public Health 2011;11:450. doi: 10.1186/1471-2458-11-450 [published Online First: 2011/06/11]

23. Eeg-Olofsson K, Cederholm J, Nilsson PM, et al. Glycemic control and cardiovascular disease in 7,454 patients with type 1 diabetes: an observational study from the Swedish National Diabetes Register (NDR). Diabetes Care 2010;33(7):1640–6. doi: 10.2337/dc10-0398 [published Online First: 2010/04/29]

24. Wettermark B, Hammar N, Fored CM, et al. The new Swedish Prescribed Drug Register--opportunities for pharmacoepidemiological research and experience from the first six months. Pharmacoepidemiol Drug Saf 2007;16(7):726–35. doi: 10.1002/pds.1294 [published Online First: 2006/08/10]

25. Wallerstedt SM, Wettermark B, Hoffmann M. The First Decade with the Swedish Prescribed Drug Register - A Systematic Review of the Output in the Scientific Literature. Basic Clin Pharmacol Toxicol 2016;119(5):464–69. doi: 10.1111/bcpt.12613 [published Online First: 2016/04/27]

26. Ludvigsson JF, Almqvist C, Bonamy AK, et al. Registers of the Swedish total population and their use in medical research. Eur J Epidemiol 2016;31(2):125–36. doi: 10.1007/s10654-016-0117-y [published Online First: 2016/01/16]

27. Mattsson K, Jönsson I, Malmqvist E, et al. Maternal smoking during pregnancy and offspring type 1 diabetes mellitus risk: accounting for HLA haplotype. Eur J Epidemiol 2015;30(3):231–8. doi: 10.1007/s10654-014-9985-1 [published Online First: 2015/01/13]

28. George L, Granath F, Johansson AL, et al. Self-reported nicotine exposure and plasma levels of cotinine in early and late pregnancy. Acta Obstet Gynecol Scand 2006;85(11):1331–7. doi: 10.1080/00016340600935433 [published Online First: 2006/11/09]

29. Örtqvist AK, Lundholm C, Kieler H, et al. Antibiotics in fetal and early life and subsequent childhood asthma: nationwide population based study with sibling analysis. BMJ 2014;349:g6979. doi: 10.1136/bmj.g6979 [published Online First: 2014/11/30]

30. Brand JS, Hiyoshi A, Cao Y, et al. Maternal smoking during pregnancy and fractures in offspring: national register based sibling comparison study. BMJ 2020;368:l7057. doi: 10.1136/bmj.l7057 [published Online First: 2020/01/31]

31. Jiang H, Fine JP, Chappell R. Semiparametric analysis of survival data with left truncation and dependent right censoring. Biometrics 2005;61(2):567–75. doi: 10.1111/j.1541-0420.2005.00335.x [published Online First: 2005/07/14]

32. Smew AI, Lundholm C, Sävendahl L, et al. Familial Coaggregation of Asthma and Type 1 Diabetes in Children. JAMA Netw Open 2020;3(3):e200834. doi: 10.1001/jamanetworkopen.2020.0834 [published Online First: 2020/03/13]

33. Bjørnvold M, Munthe-Kaas MC, Egeland T, et al. A TLR2 polymorphism is associated with type 1 diabetes and allergic asthma. Genes Immun 2009;10(2):181–7. doi: 10.1038/gene.2008.100 [published Online First: 2009/01/17]

34. Quinn PD, Rickert ME, Weibull CE, et al. Association Between Maternal Smoking During Pregnancy and Severe Mental Illness in Offspring. JAMA Psychiatry 2017;74(6):589–96. doi: 10.1001/jamapsychiatry.2017.0456 [published Online First: 2017/05/04]

35. D’Onofrio BM, Class QA, Rickert ME, et al. Translational Epidemiologic Approaches to Understanding the Consequences of Early-Life Exposures. Behav Genet 2016;46(3):315–28. doi: 10.1007/s10519-015-9769-8 [published Online First: 2015/11/23]

36. D’Onofrio BM, Class QA, Rickert ME, et al. Preterm birth and mortality and morbidity: a population-based quasi-experimental study. JAMA Psychiatry 2013;70(11):1231–40. doi: 10.1001/jamapsychiatry.2013.2107 [published Online First: 2013/09/27]

37. VanderWeele TJ, Ding P. Sensitivity Analysis in Observational Research: Introducing the E-Value. Ann Intern Med 2017;167(4):268–74. doi: 10.7326/m16-2607 [published Online First: 2017/07/12]

38. Magnus MC, Tapia G, Olsen SF, et al. Parental Smoking and Risk of Childhood-onset Type 1 Diabetes. Epidemiology 2018;29(6):848–56. doi: 10.1097/ede.0000000000000911 [published Online First: 2018/08/04]

39. Kataria Y, Gaewsky L, Ellervik C. Prenatal smoking exposure and cardio-metabolic risk factors in adulthood: a general population study and a meta-analysis. Int J Obes (Lond) 2019;43(4):763–73. doi: 10.1038/s41366-018-0206-y [published Online First: 2018/09/21]

40. Halder N, Lal G. Cholinergic System and Its Therapeutic Importance in Inflammation and Autoimmunity. Front Immunol 2021;12:660342. doi: 10.3389/fimmu.2021.660342 [published Online First: 2021/05/04]

41. Wang DW, Zhou RB, Yao YM, et al. Stimulation of α7 nicotinic acetylcholine receptor by nicotine increases suppressive capacity of naturally occurring CD4+CD25+ regulatory T cells in mice in vitro. J Pharmacol Exp Ther 2010;335(3):553–61. doi: 10.1124/jpet.110.169961 [published Online First: 2010/09/17]

42. Mabley JG, Pacher P, Southan GJ, et al. Nicotine reduces the incidence of type I diabetes in mice. J Pharmacol Exp Ther 2002;300(3):876–81. doi: 10.1124/jpet.300.3.876 [published Online First: 2002/02/28]

43. Basta PV, Basham KB, Ross WP, et al. Gestational nicotine exposure alone or in combination with ethanol down-modulates offspring immune function. Int J Immunopharmacol 2000;22(2):159–69. doi: 10.1016/s0192-0561(99)00074-0 [published Online First: 2000/02/24]

44. Zhou L, Tao X, Pang G, et al. Maternal Nicotine Exposure Alters Hippocampal Microglia Polarization and Promotes Anti-inflammatory Signaling in Juvenile Offspring in Mice. Front Pharmacol 2021;12:661304. doi: 10.3389/fphar.2021.661304 [published Online First: 2021/05/29]

45. Avşar TS, McLeod H, Jackson L. Health outcomes of smoking during pregnancy and the postpartum period: an umbrella review. BMC Pregnancy Childbirth 2021;21(1):254. doi: 10.1186/s12884-021-03729-1 [published Online First: 2021/03/28]

46. Lampousi AM, Carlsson S, Löfvenborg JE. Dietary factors and risk of islet autoimmunity and type 1 diabetes: a systematic review and meta-analysis. EBioMedicine 2021;72:103633. doi: 10.1016/j.ebiom.2021.103633 [published Online First: 2021/10/18]

47. Gaysina D, Fergusson DM, Leve LD, et al. Maternal smoking during pregnancy and offspring conduct problems: evidence from 3 independent genetically sensitive research designs. JAMA Psychiatry 2013;70(9):956–63. doi: 10.1001/jamapsychiatry.2013.127 [published Online First: 2013/07/26]

